# Coronary artery calcium mass measurement based on integrated intensity and volume fraction techniques

**DOI:** 10.1101/2023.01.12.23284482

**Authors:** Dale Black, Xingshuo Xiao, Sabee Molloi

## Abstract

**Purpose:** Agatston scoring does not detect all the calcium present in computed tomography scans of the heart. A technique that removes the need for thresholding and quantifies calcium mass more accurately and reproducibly is needed.

**Approach:** Integrated intensity and volume fraction techniques were evaluated for accurate quantification of calcium mass. Integrated intensity calcium mass, volume fraction calcium mass, Agatston scoring and spatially weighted calcium scoring were compared to known calcium mass in simulated and physical phantoms. The simulation was created to match a 320-slice CT scanner. Fat rings were added to the simulated phantoms, which resulted in small (30×20 cm^2^), medium (35×25 cm^2^), and large (40×30 cm^2^) phantoms. Three calcification inserts of different diameters and hydroxyapatite densities were placed within the phantoms. All the calcium mass measurements were repeated across different beam energies, patient sizes, insert sizes, and densities. Physical phantom images from a previously reported study were then used to evaluate the accuracy and reproducibility of the techniques.

**Results:** Both integrated intensity calcium mass and volume fraction calcium mass yielded lower root mean squared error (RMSE) and deviation (RMSD) values than Agatston scoring in all the measurements in the simulated phantoms. Specifically, integrated calcium mass (RMSE: 0.50 mg, RMSD: 0.49 mg) and volume fraction calcium mass (RMSE: 0.59 mg, RMSD: 0.58 mg) were more accurate for the low-density calcium measurements than Agatston scoring (RMSE: 3.5 mg, RMSD: 2.2 mg). Similarly, integrated calcium mass (9.72%) and volume fraction calcium mass (10.19%) had fewer false-negative (CAC=0) measurements than Agatston scoring (38.89%).

**Conclusion:** The integrated calcium mass and volume fraction calcium mass techniques can potentially improve risk stratification for patients undergoing calcium scoring and further improve risk assessment compared to Agatston scoring.

## 1. Introduction

The coronary artery calcification (CAC) score is a marker of atherosclerotic disease ^1^. CAC scoring is a test performed using computed tomography (CT) that measures the amount of calcium buildup within the walls of the coronary arteries and is an essential predictor of coronary heart disease. The leading cause of death in the United States is coronary heart disease, killing 659,000 people annually, and coronary heart disease is the third leading cause of mortality worldwide ^2^.

Agatston scoring is the most common CAC scoring technique ^3^ and is a good predictor of major adverse cardiac events (MACE) ^4^. Although, studies have shown that a significant number of patients have been characterized as having no calcium (CAC=0) while still developing MACE ^5^. This is likely due in part to the intensity thresholding requirements associated with the Agatston scoring technique, so other approaches like spatially weighted calcium scoring have been put forth as an alternative to Agatston scoring. Spatially weighted calcium scoring improves upon traditional calcium scoring by avoiding thresholding and has been shown to predict MACE more accurately ^6,7^. Spatially weighted calcium scoring is still limited in distinguishing non-zero CAC from noise. It also lacks quantitative insight as it is an arbitrary score without any direct physical association.

The Agatston technique can be used to estimate calcium volume and calcium mass. The calcium mass score acquired via the Agaston technique is fundamentally similar to traditional Agatston calcium scoring and suffers from many of the same limitations inherent to the Agatston scoring approach ^8,9^. A previous study also showed that calcium mass quantification via the Agatston approach produces up to 50% underestimation of calcium mass for large patients ^10^.

A CAC measurement technique that improves the accuracy, reproducibility, sensitivity, and specificity of the measured calcium mass would help to improve coronary heart disease diagnosis and stratify risk more accurately, especially for calcifications near the threshold of detectability. A more quantitative calcium mass calculation would likewise help improve risk stratification and diagnosis of coronary heart disease. An integrated intensity or Hounsfield unit (HU) technique recently demonstrated the ability to accurately assess coronary artery cross-sectional area near the threshold of detectability ^11^. A similar approach based on volume fractions of two materials within a voxel can be used for calcium mass quantification. These techniques have not previously been used for calcium mass quantification.

This study used the integrated HU and volume fraction mass quantification techniques to quantify coronary artery calcium in CT scans by accounting for the partial volume effect. These new calcium mass quantification techniques, integrated calcium mass, and volume fraction calcium mass were evaluated on simulated CT scans where the total calcium mass was known. These techniques, along with the standard Agatston scoring technique and the recently introduced spatially weighted calcium scoring technique, were compared to the known calcium mass to determine their accuracy and precision.

## 2. Method

### 2.1 Simulation

The simulation study was set to match the scanning parameters of the 320-slice CT scanner (Canon Aquilion One, Canon America Medical Systems, Tustin, CA), as previously reported ^12^. The X-ray spectrum was created with an interpolating polynomial model ^13^. The linear attenuation coefficients were made from their chemical composition ^14^. Poisson noise was added to simulate quantum noise. The simulation did not include Compton scatter, but beam hardening was included. A3200×2200 pixel digital phantom was designed based on an anthropomorphic thorax phantom with a size of 30×20 cm^2^ (QRM-Thorax, QRM, Möhrendorf, Germany). To simulate different patient sizes, additional fat rings emulated by a mixture of 20% water and 80% lipid were added, which resulted in a medium-sized phantom of 35×25 cm^2^ and a large-sized phantom of 40×30 cm^2^. There were nine calcification inserts within the thorax with different densities and sizes. Three calcification inserts of different diameters (1, 3, and 5 mm) and different hydroxyapatite (HA) densities were placed within each phantom. For the normal-density study, the HA densities in the inserts were 200, 400, and 800 mgHAcm^-3^. For the low-density study, the densities were changed to 25, 50, and 100 mgHAcm^-3^. Each phantom also contained a 10 mm diameter calibration rod. All phantom sizes and density levels were scanned using 80, 100, 120, and 135 kV tube voltages. For small, medium, and large patient sizes, the exposure value was adjusted to 0.9, 2.0, and 5.4 mR, respectively, resulting in similar noise levels for different-sized phantoms.

Simulation materials and geometries are shown in Figure 1. Acquisition and reconstruction parameters for the simulated and physical phantoms are shown in Table 1. The calibration rods were all 10 mm in diameter. All calcium scoring measurements were repeated across each kV, patient size, calcium insert size, and calcium insert density.

**Table 1.** Acquisition and reconstruction parameters for the simulated and physical phantoms. a. approximate detector thickness.

| Parameter | Simulation | Physical |
| --- | --- | --- |
| Manufacturer | Canon | Canon |
| CT System | Aquilion One Vision | Aquilion One Vision |
| Reconstruction | FBP | FBP |
| Tube Voltage (kV) | 80, 100, 120, 135 | 120 |
| Exposure Small (mR) | 0.9 | 15 |
| Exposure Medium (mR) | 2 | N/A |
| Exposure Large (mR) | 5.4 | 84 |
| Slice Thickness (mm) | 0.5 | 3.0 |
| Matrix Size Small (pixels) | 640 x 440 | 512 x 512 |
| Matrix Size Medium (pixels) | 740 x 540 | N/A |
| Matrix Size Large (pixels) | 840 x 640 | 512 x 512 |
| Detector Element Width (mm) | 0.5 | 0.5 |
| Detector Thickness (mm) | 0.5 | 0.5<sup>a</sup> |

**Fig. 1.**
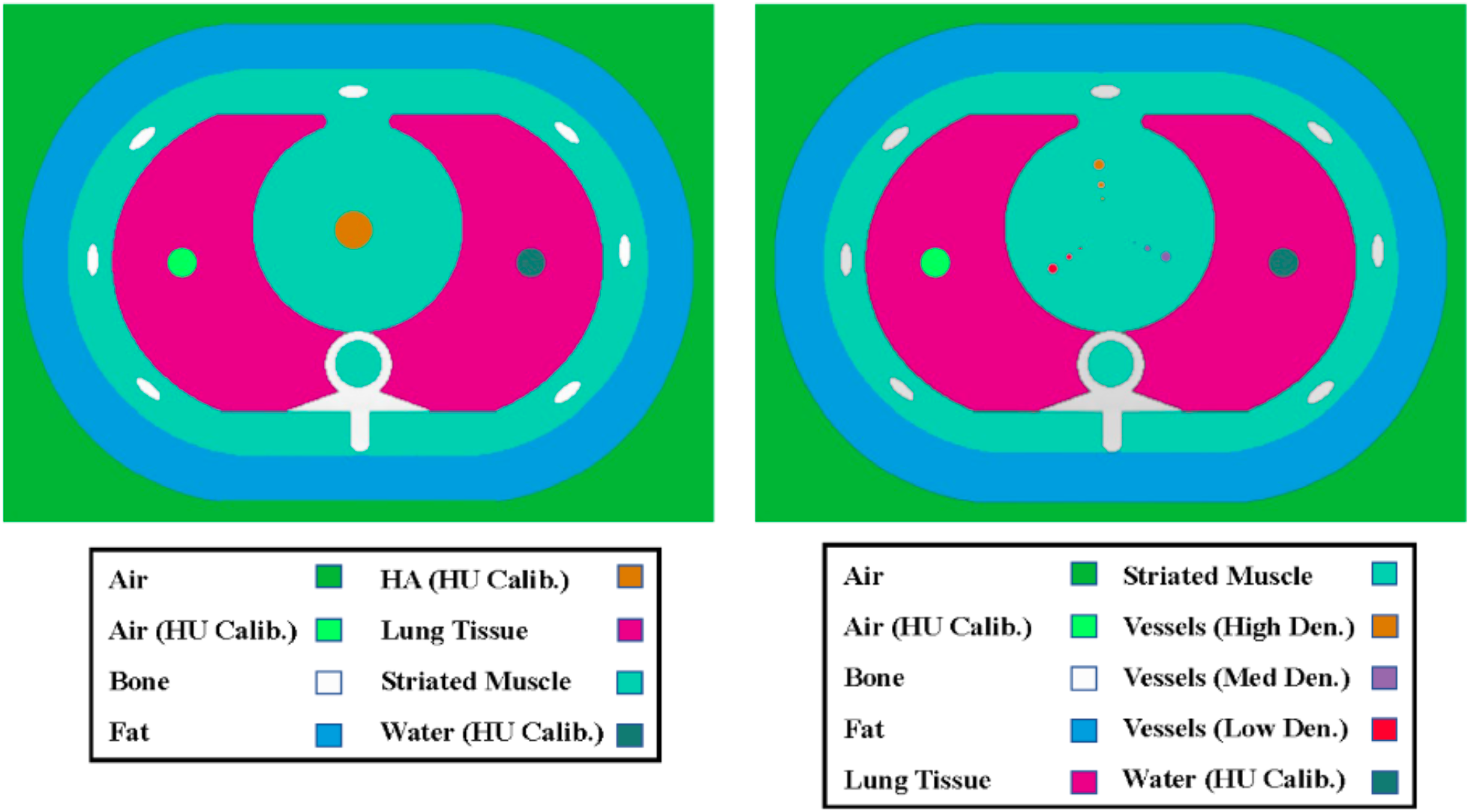
Shows a sketch of the simulated phantom with the colors highlighting the different materials in the simulated phantoms.

Segmenting regions of interest (ROIs) is important in calcium measurement. For this study, segmentations were done automatically based on previous work by Praagh et al. ^15^ and adapted for simulated phantoms. The automatic segmentation approach effectively segments calcium inserts based on the known geometry of the simulated phantom without requiring manual intervention.

To study the effect that motion has on Agatston scoring, integrated calcium mass, volume fraction calcium mass, and spatially weighted calcium scoring, we simulated motion using a previously reported random motion filter ^16,17^. We then performed repeated analysis on this simulated motion data to better understand the effect that motion has on all four calcium scoring techniques.

### 2.2 Physical phantom

This study also utilized an anthropomorphic thorax phantom (QRM-Thorax, QRM, Möhrendorf, Germany) with an insert containing calcium (Cardiac Calcification Insert (CCI), QRM, Möhrendorf, Germany). All images were acquired by Praagh et al. ^15^, and a subset of scans was included in this analysis. The cardiac calcification insert phantom consisted of nine calcification inserts made up of hydroxyapatite (HA). Within the cardiac calcification insert phantom, two calibration rods consisting of water-equivalent material and 200 mgHAcm^-3^ were also present. The calcifications had diameters and lengths of 1.0, 3.0, and 5.0 mm. Three different densities were present in the phantom for each calcification size: 200, 400, and 800 mgHAcm^-3^. Two different patient sizes, small and large, were included in this analysis by the addition of a fat ring. This fat ring increased the phantom size from 30.0 × 20.0 cm^2^ to 40.0 × 30.0 cm^2^. Four scans, two small and two large patient scans were included in this analysis. Figure 2 Shows axial slice views of various phantoms included in this study. Fig. 2A shows a simulated small phantom with normal-density inserts (200, 400, 800 mgHAcm^-3^) and a tube voltage of 120 kV. Fig. 2B shows a simulated large phantom with low-density inserts (25, 50, 100 mgHAcm^-3^) and a tube voltage of 120 kV with simulated motion. Fig. 2C shows an axial slice of the calibration rod (200 mgHAcm^-3^) for a simulated medium phantom with a tube voltage of 100 kV. Fig. 2D shows an axial slice view of a physical QRM Thorax Phantom with a Cardio Calcification Insert. The red arrow shows simulated beam hardening artifacts, and the blue arrows indicate simulated streaking artifacts caused by motion.

**Fig. 2.**
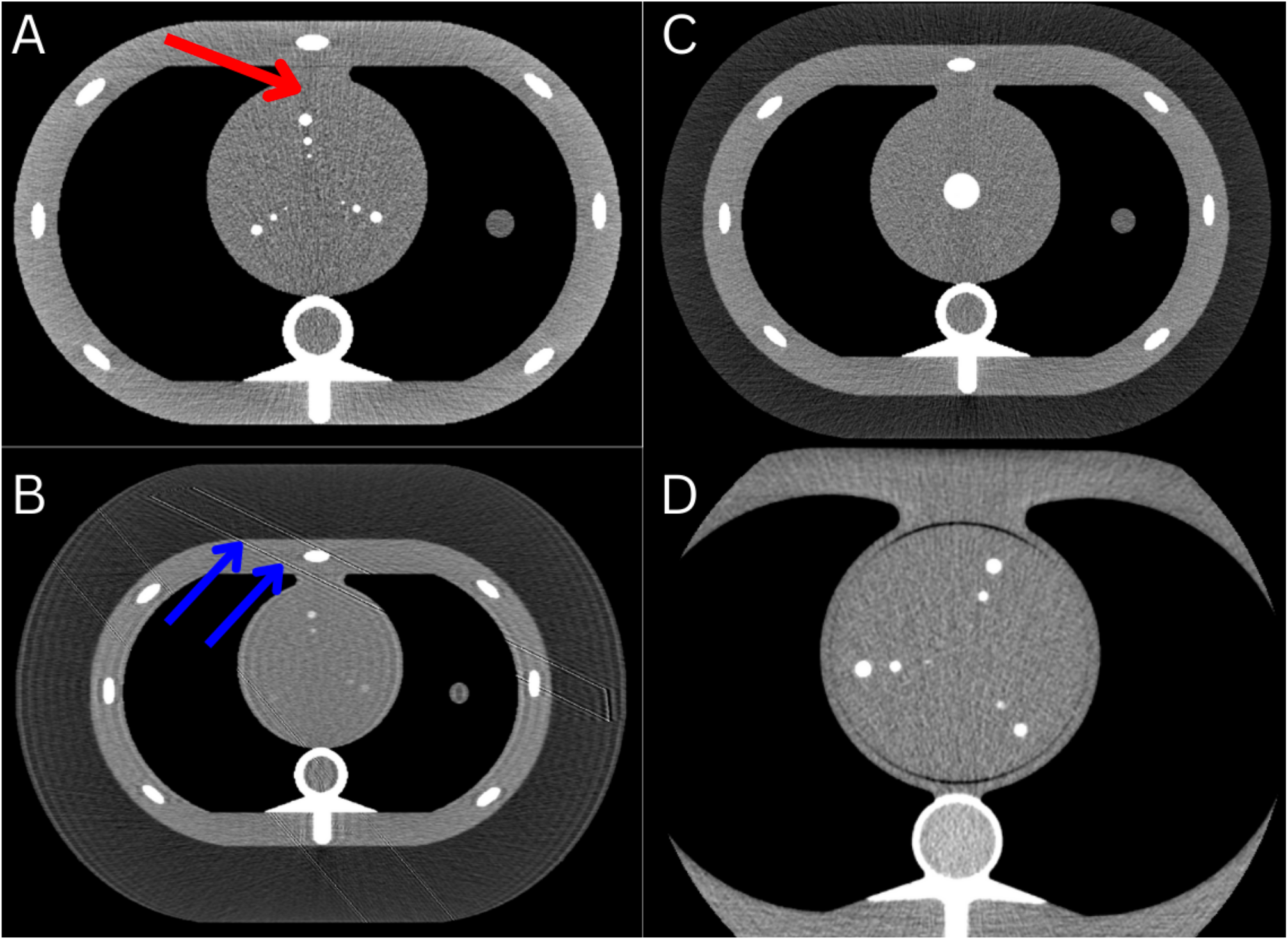
Shows axial slices of various phantoms included in this study. (A) shows a simulated small phantom with normal-density inserts (200, 400, 800 mgHAcm^-3^) and a tube voltage of 120 kV with the red arrow showing beam hardening artifacts. (B) shows a simulated large phantom with low-density inserts (25, 50, 100 mgHAcm^-3^) and a tube voltage of 120 kV with simulated motion, and the blue arrows indicate streaking artifacts caused by motion. (C) shows an axial slice of the calibration rod (200 mgHAcm^-3^) for a simulated medium phantom with a tube voltage of 100 kV. (D) shows an axial slice of a physical QRM Thorax Phantom with a Cardio Calcification Insert.

Segmenting regions of interest is an important step in calcium measurement. For this study, segmentations were done automatically based on previous work by Praagh et al. ^15^ and adapted for the Julia programming language ^18^.

Throughout this study, Agatston scoring refers to the calcium mass calculations derived from the Agatston scoring method. Spatially weighted calcium scoring refers only to the calcium score without associated physical units. Integrated calcium mass and volume fraction calcium mass refer to the calcium mass calculated via the integrated intensity approach and volume fraction mass quantification technique, respectively. This study’s calcium scoring algorithms are publicly available at https://github.com/Dale-Black/CalciumScoring.jl.

### 2.3 Agaston scoring

Agatston scoring is defined at a tube voltage of 120 kV ^3^, but recent papers have shown how Agatston scoring can be adjusted for use at lower kVs (70, 80, and 100 kV) ^19^. For this study, we assumed an exponentially decreasing trendline and extrapolated beyond to account for a higher tube voltage of 135 kV. This results in an extrapolation formula shown in Equation 1, where *y*_*thresh*_ corresponds to the extrapolated threshold (HU), and *TV* corresponds to the tube voltage (kV). All kV-specific thresholds used in this study are shown in Table 2.

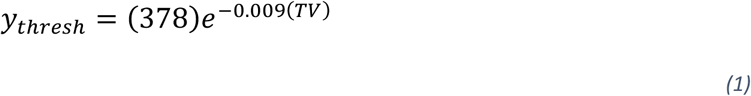

**Table 2.** Tube voltage adapted thresholds for Agatston scoring.

| Tube Voltage (kV) | Threshold (HU) |
| --- | --- |
| 80 | 177 |
| 100 | 145 |
| 120 | 130 |
| 135 | 112 |

### 2.4 Integrated calcium mass

As previously reported ^11^, the integrated Hounsfield technique assumes that although the partial volume effect influences the HU of a particular voxel, the total integrated HU within an ROI is conserved. This study addresses the issue of non-detectable CAC by adjusting the integrated HU technique for use in CAC scoring, calling it integrated calcium mass. The cross-sectional area equation (Eq. 2) can be modified for use in three-dimensional regions (Eq. 3) and applied to calcium mass quantification. Fig. 3A shows the cross-section of a simulated coronary plaque with the measurements that need to be computed for each image. *S*_*Obj*_ is the intensity (HU) of a region within the plaque containing pure calcium with no partial volume affected voxels. *S*_*Bkg*_ is the measured intensity (HU) of a ring-like section of the coronary lesion with no calcium. *I* is the sum of every voxel intensity (HU), including those voxels affected by the partial volume effect. *V* is the total number of voxels in the cross-section multiplied by the voxel size. Eq. 4 shows how to convert the volume of the object (*V*_*Obj*_) to mass (*M*_*Obj*_), where 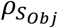 is the density of the calcification, specific to the object intensity (*S*_*Obj*_).

**Fig 3.**
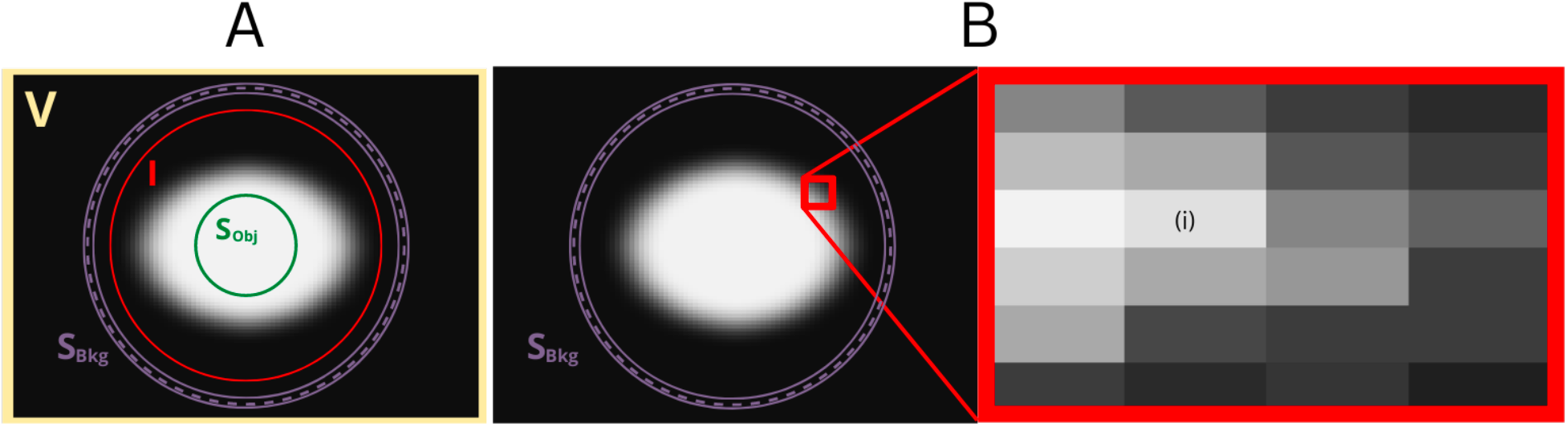
Shows two identical simulated vessel lumens with ROIs for calcium measurement. (A) Shows the ROIs needed for the integrated calcium mass technique. The central ROI that yields *S*_*Obj*_ and the background ROI that yields *S*_*Bkg*_ are unaffected by the partial volume effect, while the object ROI used to calculate *I* is affected by the partial volume effect. (B) Shows the ROIs needed for the volume fraction technique and a zoomed-in portion of the simulated vessel lumen, where *i* is the individual voxel intensity. *S*_*Bkg*_ is a ring-like background ROI unaffected by the partial volume effect. *S*_*Obj*_ is the mean intensity of known calcium density obtained from a calibration rod unaffected by the partial volume effect.

It is impractical to accurately measure the intensity of a calcification with no partial volume effect (*S*_*Obj*_) for small calcifications. Therefore, this study utilized a calibration rod and computed a volume (Eq. 3) based on the measured intensity of this calibration rod (*S*_*Obj*_), then converted the volume to mass by multiplying by the known density of that rod (Eq. 4). Integrating over the entire ROI should remove any effect from noise, assuming noise only affects individual voxels.

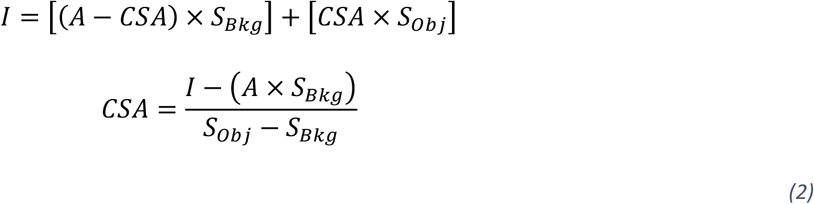

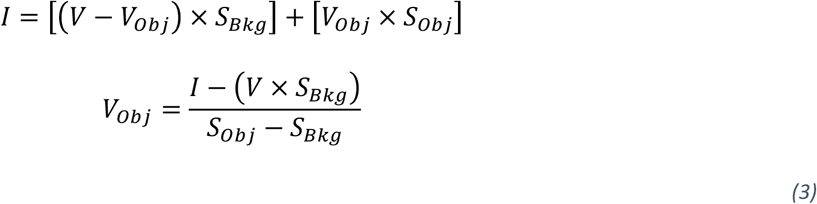

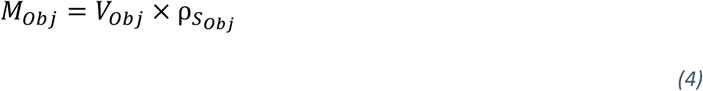

### 2.5 Volume fraction calcium mass

The volume fraction calcium mass technique is similar to integrated calcium mass, but instead of calculating the calcium within an entire ROI, the percent of calcium contained within one voxel is calculated. The percent calcium contained within each voxel is then summed up within a ROI to obtain the total percentage of calcium for that ROI (Eq. 5). Given the known size of the ROI and density of the calibration rod, the volume and mass of calcium can be calculated (Eq. 6).

Equation 5 shows how to calculate the percentage of calcium contained within each voxel. *i* is the intensity of one voxel of interest (HU), *S*_*Bkg*_ is the intensity of pure background, which can be obtained from a ring-like object as seen in Fig. 3B, and *S*_*Obj*_ is the intensity of pure calcium which can be obtained from a calibration rod of any density. The result, *k*_*i*_, is then the percentage of calcium within one voxel *i*. The entire region of voxels is then summed to give *K* which is the total percentage of calcium contained within the ROI.

Equation 6 shows how to calculate the volume of calcium (*V*_*Obj*_), given the total percentage of calcium within an ROI (*K*) and the known volume of that ROI (*V*_*ROI*_). This can then be converted into a calcium mass (*M*_*Obj*_) given the density of the calibration rod 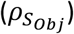. Fig. 3 shows the parameters needed to calculate mass via the integrated calcium mass technique (Fig. 3A) and the volume fraction calcium mass technique (Fig. 3B) on a simulated plaque.

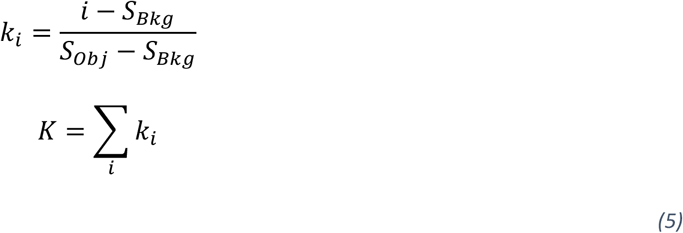

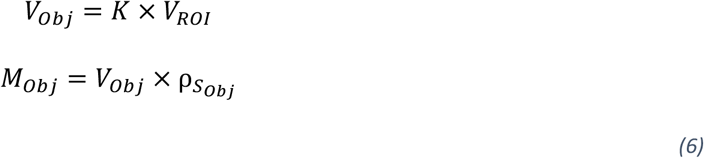

### 2.6 Spatially weighted calcium scoring

This study reimplemented the previously described spatially weighted calcium scoring technique ^6,7^ to compare integrated calcium mass and volume fraction calcium mass against a more recently proposed calcium scoring approach. The same set of voxels included in the integrated calcium mass and volume fraction calcium mass regions of interest were included in the spatially weighted calcium scoring calculations. Each voxel was assigned a weight using a normal distribution function with means and standard deviations obtained from 100 mgHAcm^-3^ calcium rod measurements across multiple simulated scans. These measurements are shown in Table 3. Only small patient sizes were used for measurements, and the signal-to-noise ratio was kept constant between different energies.

**Table 3.** Means and standard deviations of the 100 mgHAcm^-3^ calcium calibration rods.

| kV | Mean | Std |
| --- | --- | --- |
| 80 | 201.490 | 34.304 |
| 100 | 174.366 | 28.171 |
| 120 | 158.264 | 22.966 |
| 135 | 152.881 | 21.078 |

### 2.7 Statistical analysis

All calcium scoring calculations and analyses were performed using the programming language Julia ^18^. Root-mean-square error (RMSE) and root-mean-square deviation (RMSD) were calculated for all linear regression measurements to test for accuracy (RMSE) and precision (RMSD). Equation 7 shows how to calculate RMSE and RMSD. *N* is the total number of data points, ŷ is the calculated calcium masses, and *y* is either the ground truth calcium masses (RMSE) or the linear regression-based calcium masses (RMSD), which is computed based on the calculated calcium masses.

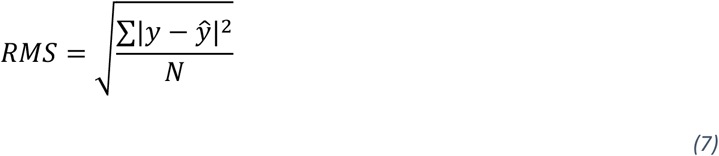

## 3. Results

### 3.1 Simulated phantoms

#### 3.1.1 Accuracy

Agatson scoring, integrated calcium mass, and volume fraction calcium mass can calculate mass directly, making the comparison between known mass and those techniques straightforward. The spatially weighted approach produces an arbitrary score correlated with the Agatston score ^6,7^. A mass calculation derived from spatially weighted calcium scoring is not practical due to the lack of thresholding in the spatially weighted technique.

Linear regression was performed for integrated calcium mass, volume fraction calcium mass, and Agatston scoring against known calcium mass on all stationary and motion-affected simulated phantoms. The low-density and normal-density phantoms were examined separately.

The correlation coefficient (r^2^), RMSE, and RMSD values were calculated for Agatston scoring, integrated calcium mass, and volume fraction calcium mass. Linear regression was performed based on the known calcium mass as the reference. Integrated calcium mass on normal-density phantoms produced an r-correlation coefficient, RMSE, and RMSD value of 1.000, 0.677 mg, and 0.602 mg, respectively. Volume fraction calcium mass on normal-density phantoms produced an r-correlation coefficient, RMSE, and RMSD value of 1.000, 0.609 mg, and 0.599 mg, respectively. Agatston scoring on normal-density phantoms produced an r-correlation coefficient, RMSE, and RMSD value of 1.000, 1.650 mg, and 0.906 mg, respectively. All normal-density accuracy measurements on the stationary phantoms are shown in Table 4.

**Table 4.**
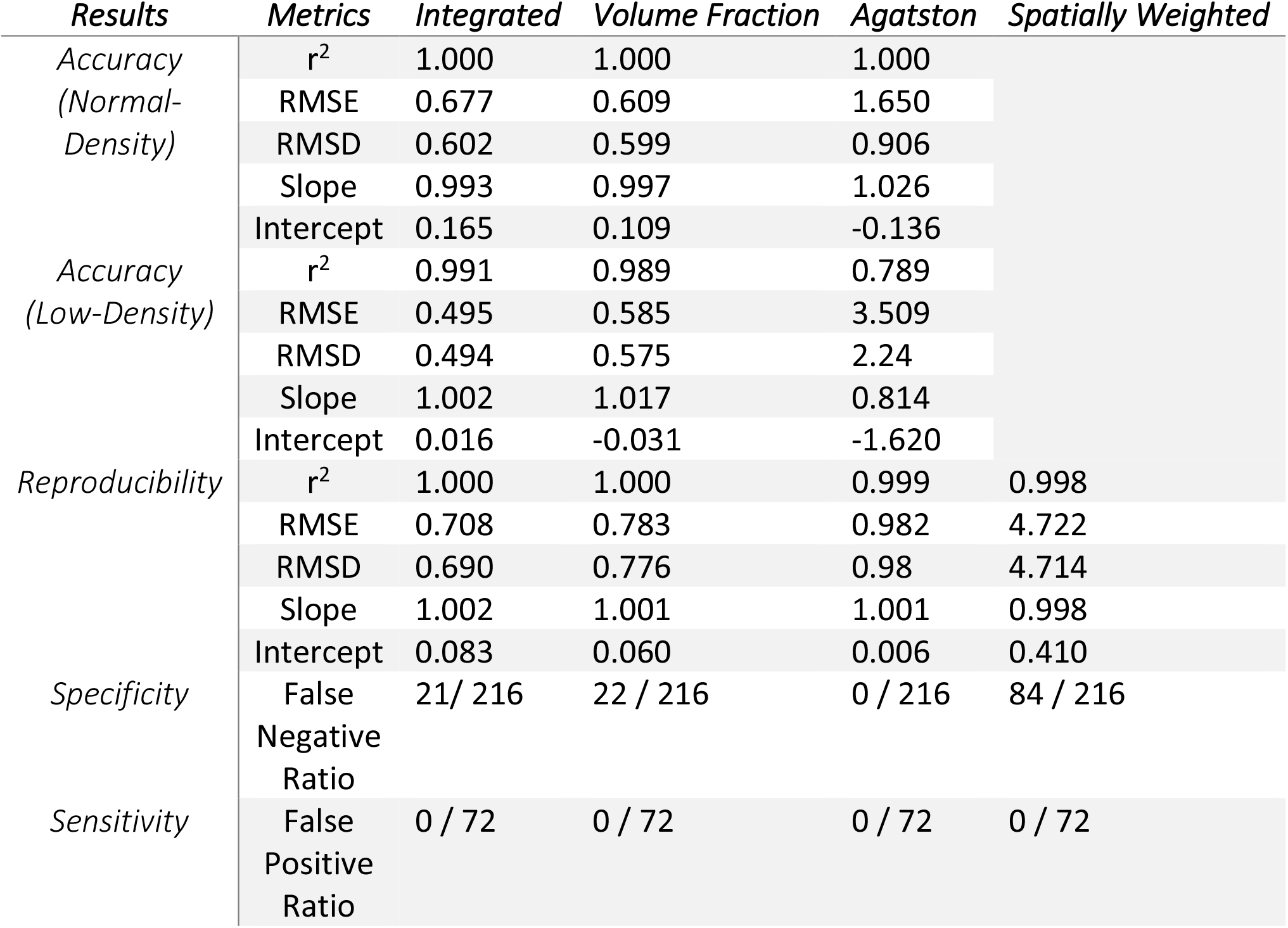
Summary of the simulated stationary phantom measurements.

Integrated calcium mass on low-density phantoms (Fig. 4A) produced an r-correlation coefficient, RMSE, and RMSD value of 0.991, 0.495 mg, and 0.494 mg, respectively. Volume fraction calcium mass on low-density phantoms (Fig. 4B) produced an r-correlation coefficient, RMSE, and RMSD value of 0.989, 0.585 mg, and 0.575 mg, respectively. Agatston scoring on low-density phantoms (Fig. 4C) produced an r-correlation coefficient, RMSE, and RMSD value of 0.789, 3.509 mg, and 2.240 mg, respectively. A similar analysis was performed for all motion-affected phantoms, which can be seen in Table 5. The trend for motion-affected data continued to show that integrated calcium mass and volume fraction calcium mass outperformed Agatston scoring for both low-density and normal-density phantoms.

**Table 5.**
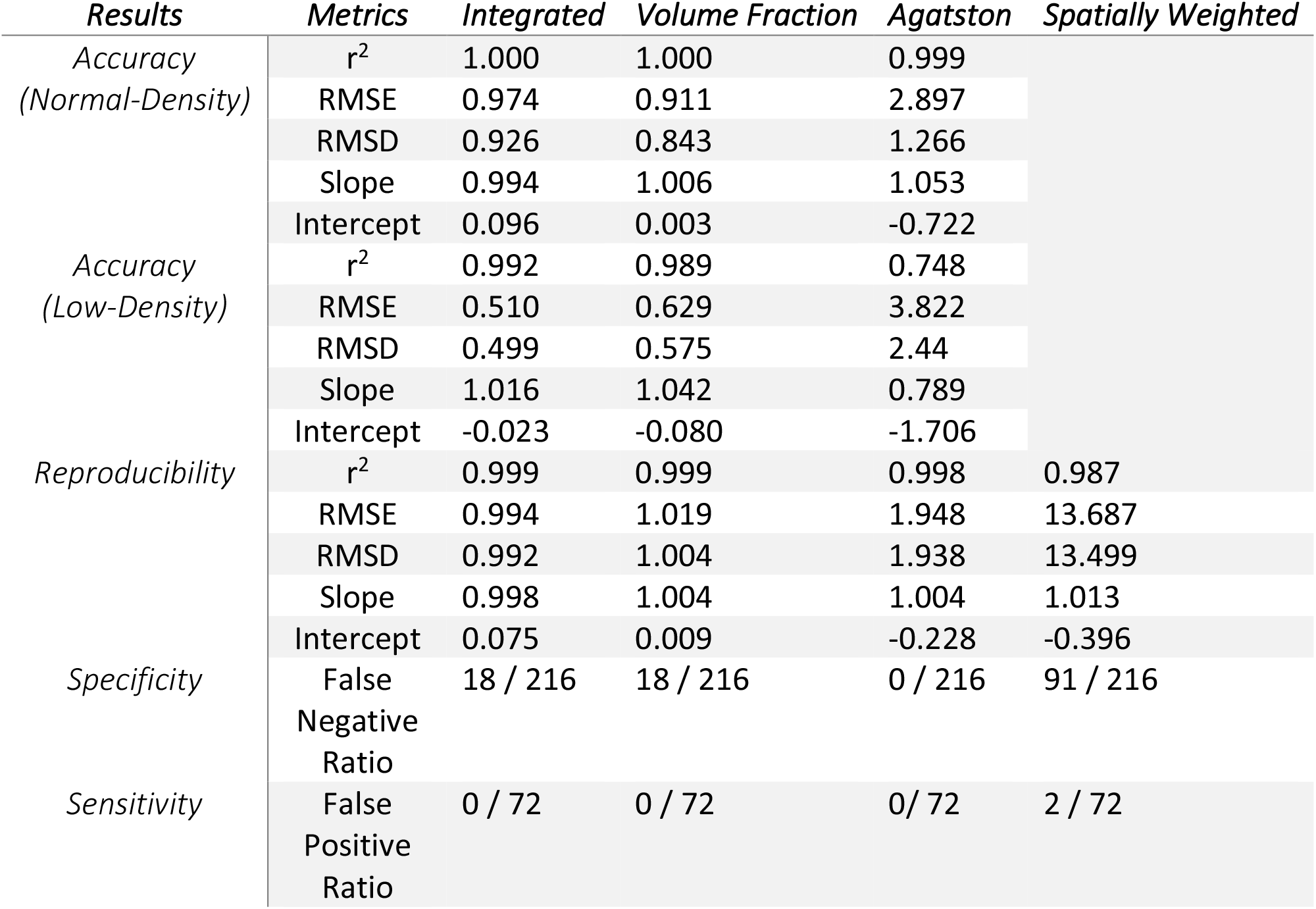
Summary of the simulated motion-affected phantom measurements.

**Fig. 4.**
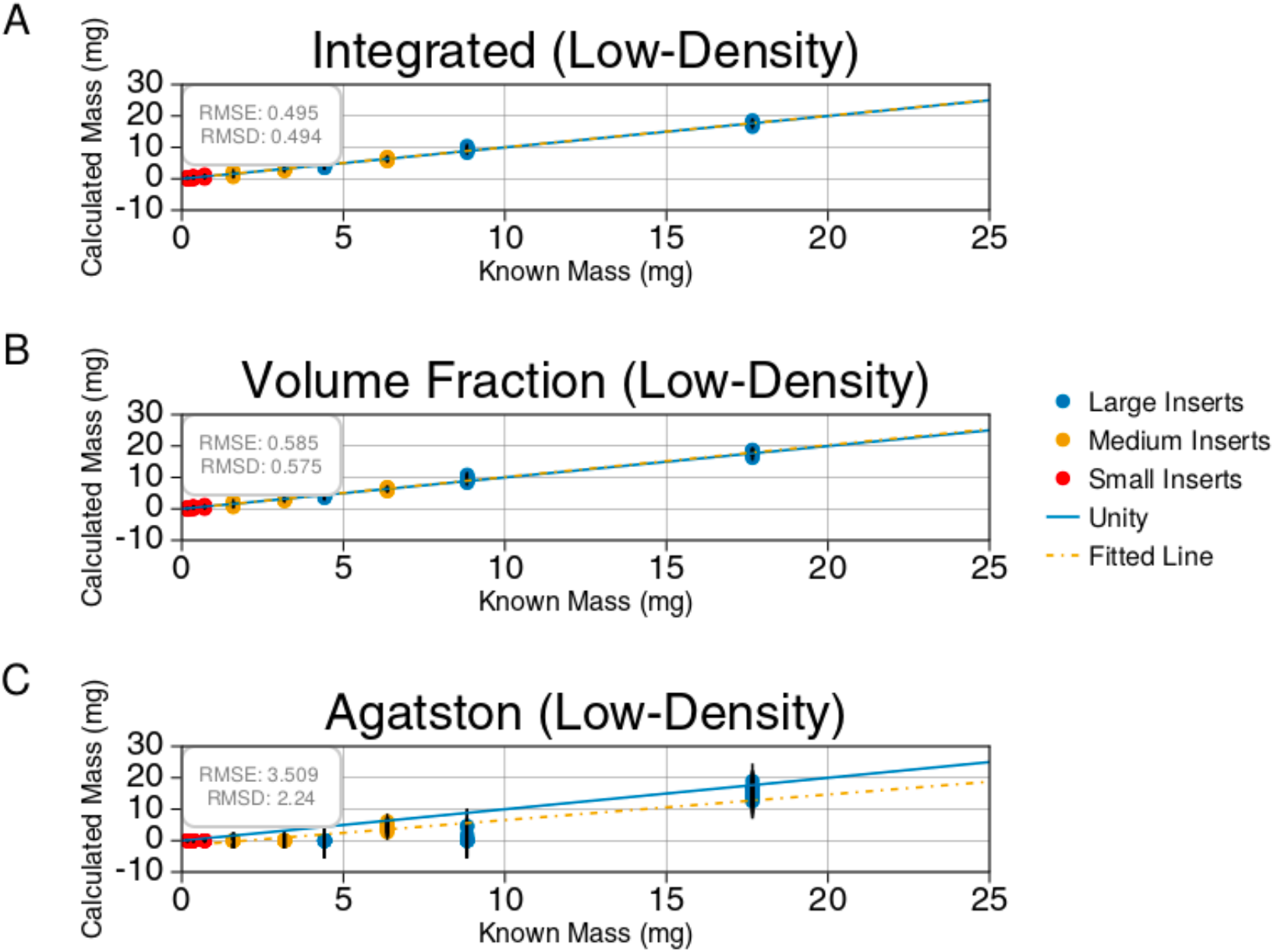
Shows the linear regression analysis comparing measured calcium to the known calcium for the low-density (25, 50, 100 mgHAcm^-3^) stationary phantoms. Every tube voltage (80, 100, 120, 135 kV) and size (small, medium, large) is included in the analysis. (A) shows the results of integrated calcium mass. (B) shows the results of the volume fraction method. (C) shows the results of Agatston mass scoring. The best fit line, along with the root mean squared error (RMSE) and root mean squared deviation (RMSD) values are shown in each plot.

#### 3.1.2 Reproducibility

This study included another set of simulated images with identical geometry to understand how repeatable the four different calcium scoring techniques are. The only variation between the two groups of images comes from the random quantum noise associated with the simulation itself. First, all false-negative values were removed from the analysis and then the reproducibility of the results was calculated. Then the results for the stationary phantoms were plotted, comparing measurements from the first set of images against the second set of images for all four scoring techniques (Fig. 5). The RMSE and RMSD values for integrated calcium mass were 0.708 mg and 0.692 mg, respectively (Fig. 5A). The RMSE and RMSD values for volume fraction calcium mass were 0.783 mg and 0.776 mg, respectively (Fig. 5B). The RMSE and RMSD values for Agatston scoring were 0.982 mg and 0.980 mg, respectively (Fig. 5C). The RMSE and RMSD values for spatially weighted calcium scoring were 4.722 mg and 4.714 mg, respectively (Fig. 5D).

**Fig. 5.**
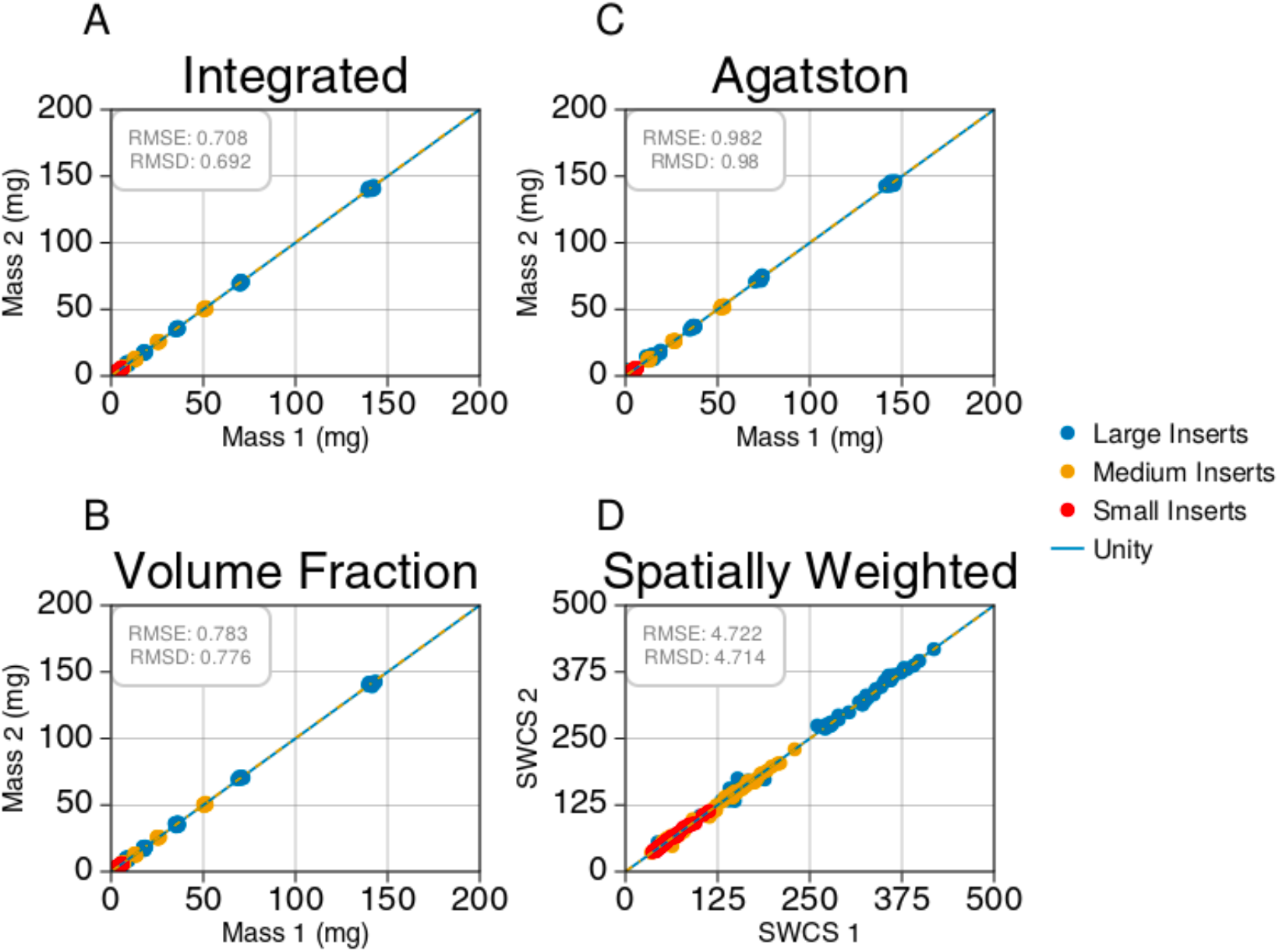
Shows reproducibility measurements for all four scoring techniques on the stationary phantoms. Every tube voltage (80, 100, 120, 135 kV), size (small, medium, large), and density (low, normal) is included in the analysis. The measurements from the first set of images were plotted against the second set of images for each technique. Integrated mass (A), Volume fraction (B), Agatston mass scoring (C), and spatially weighted calcium scoring (D) are shown along with the root mean squared error (RMSE) and root mean squared deviation (RMSD) values.

The correlation coefficient and best-fit line were also calculated for each calcium scoring technique and are shown in Table 4. The results were then repeated for the motion-affected simulated phantom data. Again, similar trends were seen in reproducibility analysis for both the stationary (Table 4) and motion-affected scans (Table 5).

#### 3.1.3 Sensitivity and specificity

The percentage of false-negative (CAC=0) and false-positive (CAC>0) scores was also calculated to understand the sensitivity and specificity of all four calcium scoring techniques. Any region containing known calcium that resulted in a CAC score of zero was determined to be a false-negative (CAC=0) score, and any region of pure background that resulted in a positive calcium score was determined to be a false-positive (CAC>0) score. This was simple to obtain for Agatston scoring, as Agatston returns zero as a possible score.

For spatially weighted calcium scoring, the score is always greater than zero, making a score of zero impossible. This is unhelpful since noise will eventually become a dominating factor for low-density and small calcifications making it difficult to distinguish between a small amount of calcium and noise, so spatially weighted calcium scores for regions of pure background were calculated across all simulated phantoms. Then the mean and standard deviation of these scores was obtained. Any spatially weighted calcium score containing regions of known calcium that resulted in a score less than the mean background score plus 1.5 standard deviations was then considered a false-negative (CAC=0) score. Likewise, any spatially weighted calcium score greater than the mean score of pure background plus 1.5 standard deviations was considered a false-positive (CAC>0) score for regions containing only background.

A similar approach for determining false-negative (CAC=0) and false-positive (CAC>0) scores was applied to integrated calcium mass and volume fraction calcium mass. Both calcium mass techniques result in physical mass calculations from negative to positive infinity. For low-density and small calcifications, noise will dominate the calcium mass results, making it important to define a threshold that can distinguish between a small amount of calcium and noise. So, a mean and standard deviation mass of background was obtained similarly to spatially weighted calcium scoring. Any integrated calcium mass or volume fraction calcium mass result containing regions of known calcium that resulted in a mass less than the mean background mass plus 1.5 standard deviations was then considered a false-negative (CAC=0) score. Likewise, integrated calcium mass or volume fraction calcium mass results greater than the mean mass of pure background plus 1.5 standard deviations was considered a false-positive (CAC>0) mass for regions containing only background.

The percentage of false-negative (CAC=0) and false-positive (CAC>0) scores were then calculated for all four techniques. For the stationary simulated phantoms, integrated calcium mass and volume fraction calcium mass techniques produced 21 and 22 false-negative (CAC=0) scores out of 216 total measurements. Spatially weighted calcium scoring produced 92 out of 216 false-negative (CAC=0) scores. Agatson scoring produced 84 false-negative (CAC=0) scores out of 216 scores. Because of the thresholding requirement applied to these calculations, zero false-positive (CAC>0) scores were produced via integrated calcium mass, volume fraction calcium mass, and spatially weighted calcium mass. Likewise, Agatston scoring produced zero false-positive (CAC>0) scores due to the 130 HU threshold and minimum connected component requirement. These results are shown in Figure 6. Results for the sensitivity and specificity of each technique, evaluated on the motion-affected phantoms, are summarized in Table 5.

**Fig 6.**
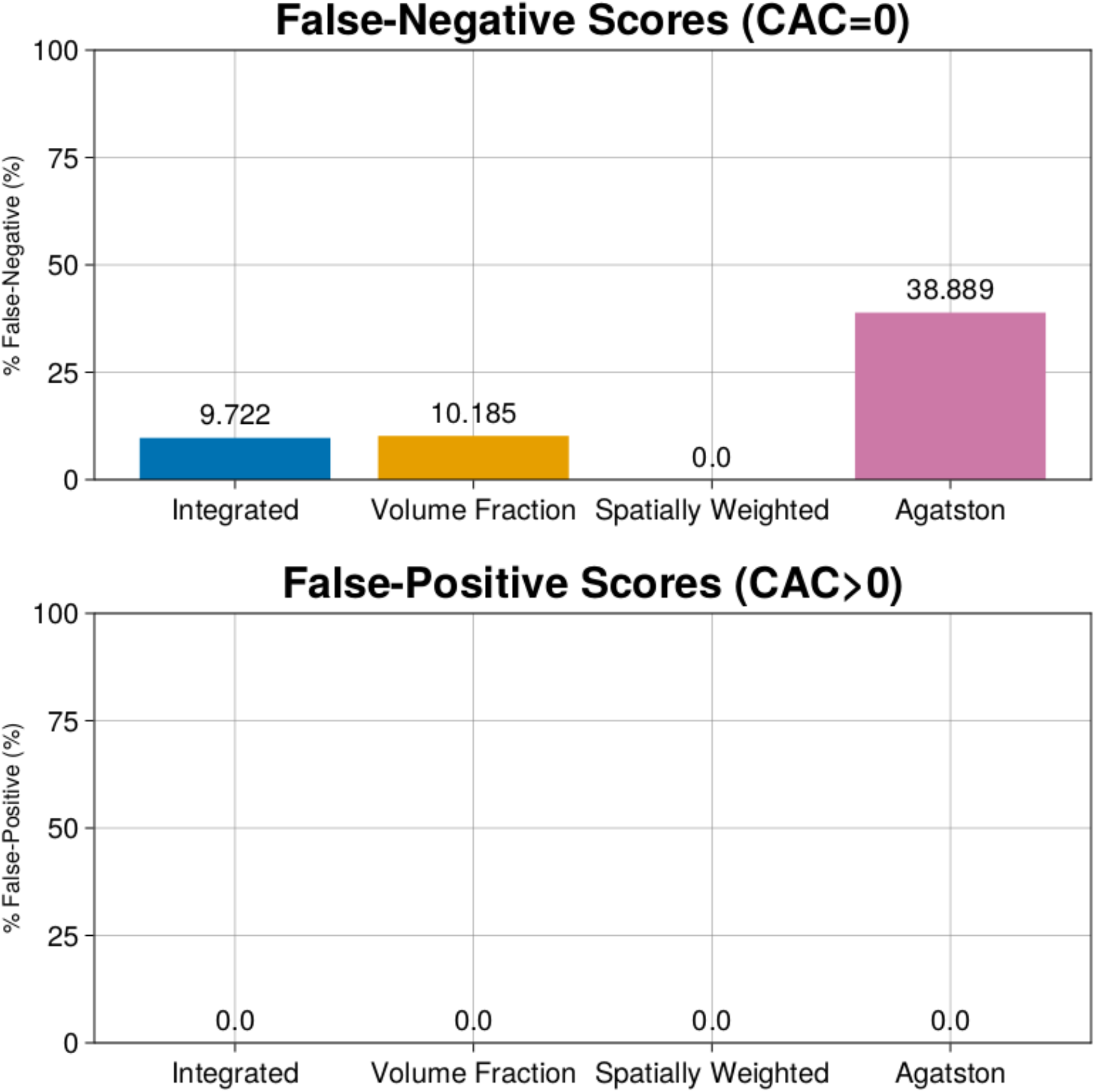
Shows the percentage of false-negative (CAC=0) and false-positive (CAC>0) scores on the stationary phantoms. Every tube voltage (80, 100, 120, 135 kV), size (small, medium, large), and density (low, normal) is included in the analysis.

### 3.2 Physical phantoms

A subset of the physical phantom scans acquired by Praagh et al. ^15^ was used for analysis. To validate the simulation results, two small and two large phantom scans acquired using the CANON scanner (Table 1) were included in this analysis. Spatially weighted calcium scoring requires a collection of 100 mgHAcm^-3^ calibration rod measurements for the weighting function. The physical QRM phantom scans only contained a 200 mgHAcm^-3^ calibration rod. Therefore, spatially weighted calcium scoring was excluded from the analysis of the physical phantom. Sensitivity and specificity results were excluded from the physical phantom analysis because the limited subset of scans was not sufficient for a proper estimation of expected false-negative (CAC=0) and false-positive (CAC>0) ratios.

#### 3.2.1 Accuracy

Linear regression was performed for integrated calcium mass, volume fraction calcium mass, and Agatston scoring against known calcium mass on all four physical phantom scans. Only normal-density (200, 400, 800 mgHAcm^-3^) inserts are included in the physical QRM phantom.

The correlation coefficient (r^2^), RMSE, and RMSD values were calculated for Agatston scoring, integrated calcium mass, and volume fraction calcium mass. Linear regression used the known calcium mass as the reference. Integrated calcium mass on the normal-density physical phantom (Fig. 7A) produced an r-correlation coefficient, RMSE, and RMSD value of 0.981, 8.811 mg, and 5.752 mg, respectively. Volume fraction calcium mass on the normal-density physical phantom (Fig. 7B) produced an r-correlation coefficient, RMSE, and RMSD value of 0.975, 8.559 mg, and 6.647 mg, respectively. Agatston scoring on the normal-density physical phantom (Fig. 7C) produced an r-correlation coefficient, RMSE, and RMSD value of 0.973, 22.485 mg, and 4.653 mg, respectively. All normal-density physical phantom accuracy measurements, including the best-fit line and the r-correlation coefficient, are shown in Table 6.

**Table 6.** Summary of the simulated motion-affected phantom measurements.

| *Results* | *Metrics* | *Integrated* | *Volume Fraction* | *Agatston* |
| --- | --- | --- | --- | --- |
| *Accuracy (Normal-Density)* | $r^2$ | 0.981 | 0.975 | 0.973 |
|  | RMSE | 8.811 | 8.559 | 22.485 |
|  | RMSD | 5.752 | 6.647 | 4.653 |
|  | Slope | 0.962 | 0.966 | 0.662 |
|  | Intercept | -5.000 | -3.907 | -3.643 |
| *Reproducibility* | $r^2$ | 0.998 | 0.998 | 0.996 |
|  | RMSE | 2.838 | 3.203 | 2.144 |
|  | RMSD | 1.906 | 1.777 | 1.792 |
|  | Slope | 1.032 | 1.041 | 1.020 |
|  | Intercept | 0.591 | 0.669 | 0.480 |
| *Specificity* | $r^2$ | 6 / 36 | 6 / 36 | 7 / 36 |
| *Sensitivity* | RMSE | 1 / 12 | 1 / 12 | 0 / 12 |

**Fig. 7.**
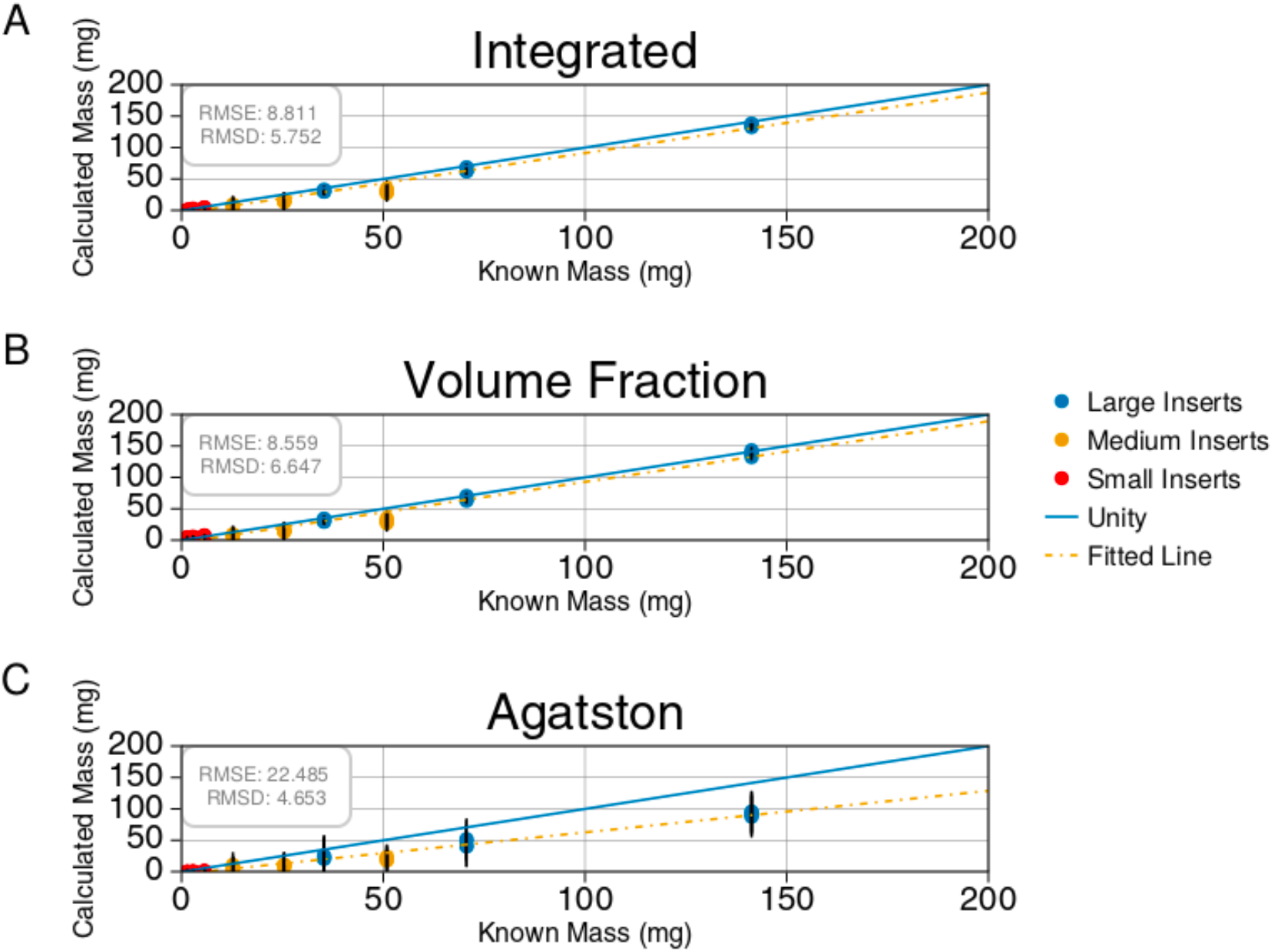
Shows the linear regression analysis comparing measured calcium to the known calcium for the normal density (200, 400, 800 mg/ cm3) physical phantom scans at a tube voltage of 120 kV. (A) shows the results of integrated calcium mass. (B) shows the results of the volume fraction method. (C) shows the results of Agatston mass scoring. The best fit line, along with the root mean squared error (RMSE) and root mean squared deviation (RMSD) values, are shown in each plot.

#### 3.2.2 Reproducibility

To analyze the reproducibility of integrated calcium mass, volume fraction calcium mass, and Agatston scoring on the physical phantom, the first small and large phantom scans were compared to the second small and large phantom scans. These scans were acquired under identical settings, with the only variation coming from a rotation of the phantom. All false-negative (CAC=0) values were removed from the analysis, and then the reproducibility of each technique was calculated.

The RMSE and RMSD values for integrated calcium mass were 2.838 mg and 1.906 mg, respectively (Fig. 8A). The RMSE and RMSD values for volume fraction calcium mass were 3.203 mg and 1.777 mg, respectively (Fig. 8B). The RMSE and RMSD values for Agatston scoring were 2.144 mg and 1.792 mg, respectively (Fig. 8C).

**Fig. 8.**
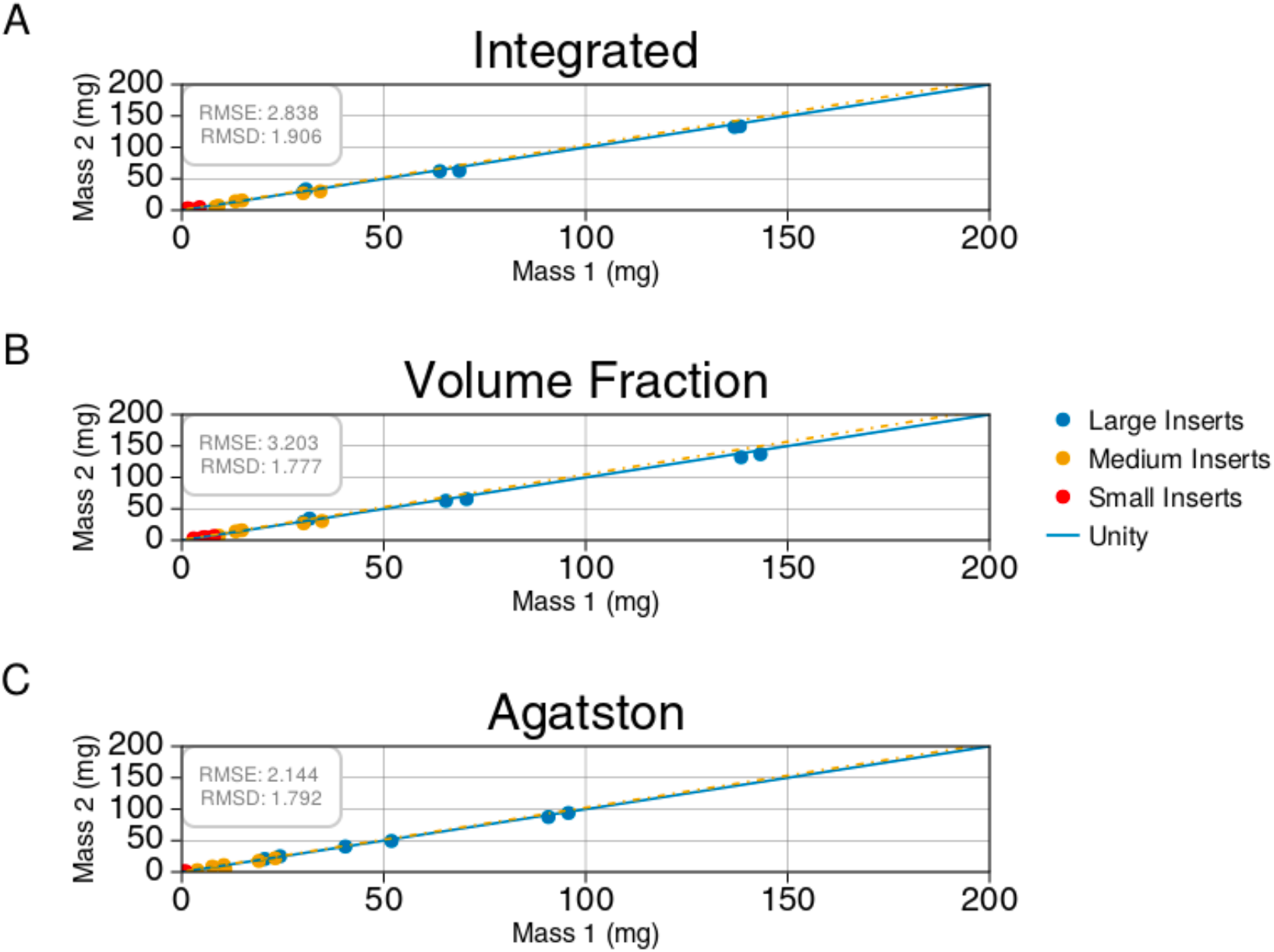
Shows reproducibility measurements on the physical phantom scans at a tube voltage of 120 kV. Measurements from two scans were plotted against a second set of measurements from two different scans. Integrated mass (A), Volume fraction (B), Agatston mass scoring (C) are shown along with the root mean squared error (RMSE) and root mean squared deviation (RMSD) values.

The up to 50% underestimation for the Agatston scoring masses (Fig. 7C) means the calculated masses are, on average, smaller than the calculated masses from the integrated and volume fraction calcium mass techniques. Since RMS values are sensitive to the magnitude of the inputs, the smaller RMSE and RMSD values for Agatston scoring, relative to integrated calcium mass and volume fraction calcium mass techniques, are explained by the underestimated mass calculations of the Agatston technique. The correlation coefficient and best-fit line were also calculated for each calcium scoring technique and are shown in Table 6.

## 4. Discussion

Calcium mass was measured on phantoms with different patient sizes, kVs, calcium sizes, and calcium densities, with and without motion. These results were then validated in a subset of physical phantom scans, acquired by Praagh et al. ^15^. The calculated calcium mass was compared against the known mass of the calcium inserts. The results indicate that integrated calcium mass and volume fraction calcium mass are more accurate, reproducible, sensitive, and specific than Agatston scoring (Tables 4, 5, and 6).

Agatston scoring has commonly been used in the past for predicting patient outcomes. However, a limitation of Agatston scoring is that it’s only defined at 120 kV, and a threshold of 130 HU is commonly used for calcium detection. However, the calcium attenuation coefficient is energy dependent, which makes scoring challenging when images are acquired at lower kVs, to reduce patient radiation dose. Recent reports have introduced correction factors for calcium measurements at lower kVs ^20,21^. Another limitation of a thresholding approach for calcium measurement is that it is affected by partial volume effect and motion. This paper introduces two new methods for calcium mass quantification based on integrated intensity (HU) and volume fraction that address the above limitations.

Previous studies have shown that up to 5% of patients with suspected zero CAC (CAC=0) within a ten-year follow up period will experience MACE, despite no detectible calcium by Agatston scoring ^22^. One question arises whether these patients had undetectable calcium by traditional Agatston scoring or simply no calcium. Integrated calcium mass, volume fraction calcium mass, and spatially weighted calcium scoring attempt to address this concern by removing the intensity thresholding requirements of standard Agatston scoring. This study shows that integrated calcium mass and volume fraction calcium mass are more sensitive to low-density calcifications than spatially weighted calcium scoring and Agatston scoring. The results showed that the percentage of false-negative (CAC=0) scores on the stationary simulated phantom were 9.722, 10.185, 0.0, and 38.889 for integrated calcium mass, volume fraction calcium mass, spatially weighted calcium scoring, and Agatston scoring, respectively (Figure 6). Compared to the existing techniques, the substantial reduction in false-negative (CAC=0) masses for the integrated calcium mass and volume fraction calcium mass techniques will help address the current limitation for patients with false-negative (CAC=0) scores.

The integrated calcium mass and volume fraction calcium mass techniques can detect calcifications that are currently undetectable by the Agatston scoring approach due to its thresholding requirement. Furthermore, a previous study has shown that calcium volume was positively and independently associated with MACE risk, and calcium density was inversely associated with MACE risk ^23^. Another study has shown that calcium density score was the strongest positive independent predictor of major adverse cardiac events, compared to Agatston score, mass score, and volume score ^4^. Disagreements between these studies are likely related to the thresholding approach of Agatston scoring, and poor reproducibility of Agatston scoring, which is also a limitation of all the traditional calcium (mass, volume, and density) scoring approaches based on the Agatston technique. Integrated calcium mass and volume fraction calcium mass provide more accurate, reproducible, and quantitative approaches to calcium measurement. Future studies on patient data comparing Agatston scoring with integrated calcium mass and volume fraction calcium mass might help explain these seemingly contradictory results better.

The integrated calcium mass and volume fraction calcium mass techniques can detect calcifications that are near the threshold of visual detectability. This is important because much of this study is focused on the accurate measurement of calcium mass for inserts near the threshold of visual detectability. In patients, coronary artery centerline estimates can be extracted in non-contrast CT scans by automatic methods, such as atlas-based approaches ^24,25^. Recently, deep learning has shown promise in automatically segmenting cardiac anatomy and has the potential to accurately segment coronary artery centerlines in non-contrast CT scans using a supervised learning approach in patient images like the OrCaScore dataset ^26^. The coronary artery centerlines can then automatically generate ROIs for calcium mass measurement of coronary artery calcification.

When acquiring the mass of calcium in the QRM phantom using Agatston scoring, it has been shown that the mass scores can be significantly different from the known physical mass ^27^. Our results (Figures 4 and 7) show that integrated calcium mass and volume fraction calcium mass measure calcium mass more accurately than Agatston scoring, on both simulated and physical phantoms. We also see an improvement in reproducibility when comparing integrated calcium mass and volume fraction calcium mass against Agatston scoring and spatially weighted calcium scoring (Fig. 5).

Although many parameters were considered in the simulation, only one type of reconstruction was utilized, which prevented us from addressing the effect that reconstruction type has on these calcium scoring techniques. Previous studies have shown good agreement between Agatston scores based on filtered back-projection, hybrid iterative reconstruction, and deep learning-based reconstruction ^28^.

Slice thickness plays an important role in calcium scoring, and traditional Agatston scoring is only defined at a slice thickness of 3 mm. Recent studies have shown that the accuracy and sensitivity of Agatston scoring are improved when slice thickness is decreased ^29^. Our simulation was limited to 0.5 mm slice thickness which is expected to provide more accurate and sensitive comparisons for Agatston scoring. Nonetheless, future studies might provide insights by varying the slice thickness. This study was also limited by the lack of realistic cardiac hardware, such as stents, known to cause blooming or motion artifacts ^30^. Future studies should account for this by including physical phantoms with realistic cardiac hardware or a robust patient data set.

Fewer total scans were included in the physical phantom analysis compared to the simulated analysis, resulting in fewer calcium scores. This is a limitation and becomes more pronounced in the sensitivity and specificity analysis, where false-negative (CAC=0) and false-positive (CAC>0) results are rare. Therefore, sensitivity and specificity analysis was not performed on the physical phantoms. In addition, motion was not included in the physical phantom analysis. More physical phantom scans, including scans affected by motion, need to be included in future studies. This would provide more robust accuracy, reproducibility, sensitivity, and specificity measurements and would result in a more reliable expected percentage of false-negative (CAC=0) and false-positive (CAC>0) scores for each calcium quantification technique.

Previous studies show that Agatston scoring consistently underestimates calcium density and volume, with even further underestimation for low-density and motion-affected plaques ^31^. Werf et al. indicate that low-density calcifications might fall below the 130 HU threshold because of blurring from motion which artificially reduces the Agatston score ^32^. Our study is consistent with these results (Fig. 4C and 7C); we showed that Agatston scoring produced the most false-negative (CAC=0) classifications for the simulated data and the physical phantom scans. Future studies are warranted in physical phantoms with lower-density calcification inserts (< 200 mgHAcm^-3^) to understand how integrated calcium mass and volume fraction calcium mass compares to Agatston mass within the low-density regime on physical data. Very high coronary artery calcium density (> 1000 mgHAcm^-3^) is quite rare, and Agatston scoring has already been shown to be a good predictor of cardiovascular disease within this subset of patients^33^.

Tzolos et al. showed that Agatston scoring struggles to detect small calcifications in the coronary arteries of patients due to the threshold requirement of 130 HU and the minimum connected component requirement of 1 mm ^34^. Our results are consistent with this study and indicate that the size of the calcium insert is a critical variable in accounting for false-negative (CAC=0) Agatston scores. The small inserts resulted in 40 false-negative (CAC=0) scores out of 216 total scores, whereas the medium and large inserts accounted for only 24 and 20 false-negative (CAC=0) scores, respectively. Density was also an important factor. Low-density calcifications resulted in 40 false-negative (CAC=0) scores out of 216 total scores, whereas the medium-density and high-density calcifications resulted in 32 and 12 false-negative (CAC=0) scores, respectively. Based on our results, integrated calcium mass and volume fraction calcium mass improved upon many of the issues associated with Agatston scoring and resulted in fewer false-negative (CAC=0) scores.

## 5. Conclusion

This study shows that both the integrated calcium mass and volume fraction calcium mass techniques improve the sensitivity, accuracy, and reproducibility of calcium mass measurement as compared to traditional Agatston scoring. This improvement in calcium scoring can potentially improve risk stratification for patients undergoing calcium scoring and further improve outcomes compared to Agatston scoring.

## Data Availability

https://github.com/Dale-Black/CalciumScoring.jl

## Acknowledgements

The authors would like to thank Drs. Martin Willemink and GD van Praagh for sharing their data for this study. A grant from Canon Medical Systems, USA, partially supported this study.

